# Local patterns of spread of influenza A(H3N2) virus in coastal Kenya over a one-year period revealed through virus sequence data

**DOI:** 10.1101/2021.09.08.21263309

**Authors:** D. Collins Owuor, Joyce M. Ngoi, Festus M. Nyasimi, Nickson Murunga, Joyce U. Nyiro, Rebecca Garten, John R. Barnes, Sandra S. Chaves, D. James Nokes, Charles N. Agoti

## Abstract

**Background:** The patterns of spread of influenza A viruses in local populations in tropical and sub-tropical regions are unclear due to sparsity of representative spatiotemporal sequence data.

**Methods:** We sequenced and analyzed 58 influenza A(H3N2) virus genomes sampled between December 2015 and December 2016 from nine health facilities within the Kilifi Health and Demographic Surveillance System (KHDSS), a predominantly rural region, covering approximately 891 km^2^ along the Kenyan coastline. The genomes were compared with 1,571 contemporaneous global sequences from 75 countries.

**Results:** We observed at least five independent introductions of A(H3N2) viruses into the region during the one-year period, with the importations originating from Africa, Europe, and North America. We also inferred 23 virus location transition events between the nine facilities included in the study. International virus imports into the study area were captured at the facilities of Chasimba, Matsangoni, Mtondia, and Mavueni, while all four exports from the region were captured from the Chasimba facility, all occurring to Africa destinations. A strong spatial clustering of virus strains at all locations was observed associated with local evolution.

**Conclusion:** Our study shows that influenza A(H3N2) virus epidemics in local populations appear to be characterized by limited introductions followed by significant local spread and evolution.

## INTRODUCTION

Two subtypes of human influenza type A virus (IAV), A(H3N2) and A(H1N1)pdm09, and two lineages of human influenza type B virus (IBV), Victoria and Yamagata, currently co-circulate in human populations [1] causing annual seasonal epidemics globally [2-6]. These viruses belong to the family *Orthomyxoviridae*, which are enveloped, negative-sense, single-stranded RNA viruses with segmented genomes [7]. IAVs evolve rapidly and undergo immune driven selection through accumulation of amino acid changes, especially at antigenic sites of hemagglutinin (HA) glycoproteins [8-10]. These amino acid sequence drifts on HA are observed more frequently in A(H3N2) virus than A(H1N1)pdm09 virus [11, 12]. Since 2009, antigenic drift of A(H3N2) viruses has resulted in emergence of several genetic groups (i.e., clades, subclades, and subgroups) globally, for example, clade 1 to 7 viruses [13]. Clade 3 viruses are the most genetically diverse and are divided into several genetic groups: subclades 3A, 3B, and 3C; and subgroups 3C.1, 3C.2, 3C.2a, 3C.3, and 3C.3a. We recently characterized A(H3N2) viruses that circulated in coastal Kenya between 2009 and 2017 and revealed co-circulation of multiple A(H3N2) virus genetic groups among hospitalized patients and outpatients [5].

While the global spread of seasonal influenza viruses has been studied intensively using phylogenetic and phylogeographic approaches [14-20], their local patterns of spread remain less clear [21], especially at city-wide or town scale. Although it is important to understand how diseases spread around the globe, local spread is most often the main driver of novel cases of respiratory diseases such as COVID-19 or influenza [21]. Transmission patterns have been recorded at different levels of human social clustering. Household studies have been performed to investigate person-to-person influenza transmission [22]. Studies of college campuses using phylogenetic methods have revealed extensive mixing of influenza virus strains among college students [23]. City-wide and countrywide, transmission of influenza in a season is characterized by majorly multiple virus introductions into cities [21] and countries [24-26]; viruses then spread from multiple geographical locations to multiple geographical destinations following introduction [24-26]. However, there is a paucity of studies that describe the patterns of spread of influenza on a local scale (city or town) due to sparsity of representative spatiotemporal sequence data from defined sub-populations residing in the same geography within a country [21, 27].

Given this gap in knowledge, we studied the patterns of spread of A(H3N2) virus in coastal Kenya using virus next generation sequencing (NGS) data collected over a one-year period between December 2015 and December 2016, along with 1,571 global A(H3N2) virus sequence data from 75 countries to provide a phylogenetic context.

## MATERIALS AND METHODS

### Study design

The samples analysed in this study were collected from nine health facilities within the Kilifi Health and Demographic Surveillance System (KHDSS) [28, 29]. The KHDSS, in Kilifi County, is a predominantly rural location along the Kenyan coastline covering an area of 891 km^2^; **Figure 1, Panel A**. The KHDSS monitors a population of approximately 296,000 residents (2016 census) through household enumeration visits conducted every 4 months [29]. The KHDSS area has 21 public health facilities (including Kilifi County Hospital (KCH)) that receives outpatients and operates under the Kenya Ministry of Health. In total, nine of these facilities, spread throughout the KHDSS, were selected for this study: Matsangoni, Ngerenya, Mtondia, Sokoke, Mavueni, Jaribuni, Chasimba, Pingilikani, and Junju, **Figure 1, Panel A**. Non-residents and residents of the KHDSS presenting to these facilities were included. A total of 5,796 nasopharyngeal (NP) swab samples were taken from outpatients across all age groups presenting with acute respiratory illness (ARI). A comprehensive description of the study area and ARI surveillance at the KHDSS outpatient health facilities has been provided in our previous publications [28, 29].

**Figure 1.**
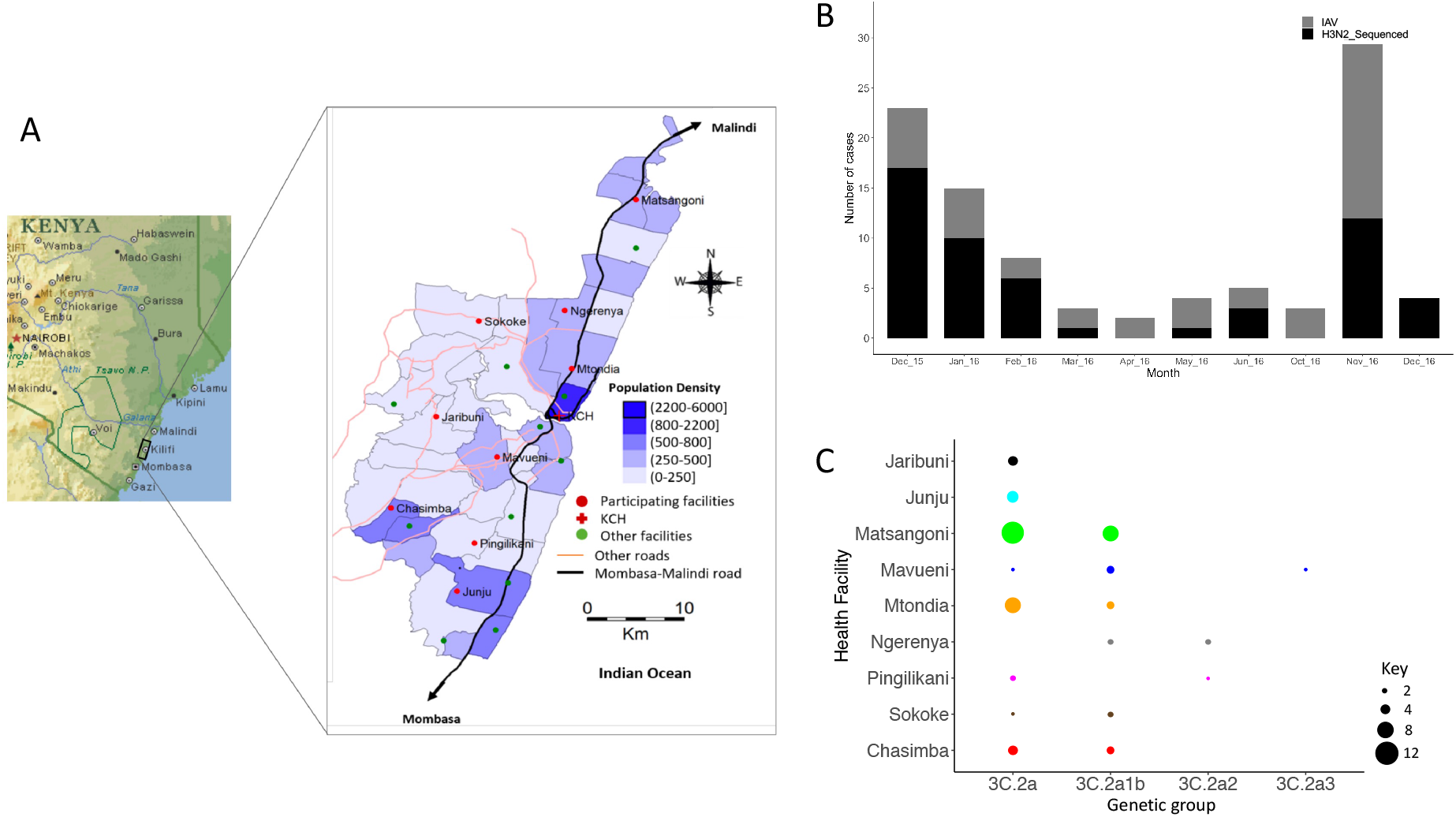
Study locations, A(H3N2) virus detections in the enrolled health facilities, and distribution of detected A(H3N2) virus genetic groups by health facility in Kilifi, Kenya. **A** Map of the KHDSS area in Kilifi, coastal Kenya showing the spatial distribution of the enrolled KHDSS health facilities (from [29]; adapted courtesy of Creative Commons Attribution Licensing). **B** Bar plot showing number of IAV virus positive samples and sequenced A(H3N2) virus positive samples by month in Kilifi between December 2015 and December 2016. All collected IAV positive samples and sequenced A(H3N2) virus samples are indicated by color (all positive IAV samples in gray – IAV; sequenced samples in black – H3N2_Sequenced) as shown in the color key. **C** Bubble plot showing A(H3N2) virus genetic groups distributed by KHDSS health facility. The size of the circle is proportional to number of samples as shown in the counts key below the figure. KHDSS, Kilifi Health and Demographic Surveillance System; IAV, influenza A virus.

### Sample handling and molecular screening

NP samples were stored in viral transport medium (VTM) at -80°C prior to molecular screening and subsequent laboratory processing [29, 30]. Samples were screened for a range of respiratory viruses, including IAV, using a multiplex (MPX) reverse transcription (RT)-PCR assay employing Qiagen QuantiFast multiplex RT-PCR kit (Qiagen); a real-time PCR cycle threshold (Ct) of <35.0 was used to define virus-positive samples [30]. A total of 97 IAV positive specimens were available for genome sequencing from the KHDSS facilities.

### RNA extraction and multi-segment real-time PCR (M-RTPCR)

Viral nucleic acid extraction from IAV positive samples was performed using QIAamp Viral RNA Mini Kit (Qiagen). Extracted RNA was reverse transcribed, and the entire IAV genome amplified in a single M-RTPCR reaction using the Uni/Inf primer set [31]. Successful amplification was evaluated by running the products on a 2% agarose gel and visualized on a UV transilluminator after staining with RedSafe Nucleic Acid Staining solution (iNtRON Biotechnology Inc.,).

### IAV next generation sequencing and genome assembly

Following M-RTPCR, amplicon purification and library preparation was conducted as previously described [5]. Libraries were then diluted to 2 nmol/L, pooled, denatured, diluted to 12.5 pmol/L and sequenced on the Illumina MiSeq using 2 × 250 bp paired-end reads using the MiSeq v2 500 cycle kit with 5% Phi-X (Illumina) spike-in. Sequence assembly was performed using the Iterative Refinement Meta-Assembler (IRMA) [32] in EDGE Bioinformatics environment [33] using IRMA default settings: median read Q-score filter of 30; minimum read length of 125; frequency threshold for insertion and deletion refinement of 0.25 and 0.6, respectively; Smith-Waterman mismatch penalty of 5; and gap opening penalty of 10. All generated sequence data were deposited in the Global Initiative on Sharing All Influenza Data (GISAID) EpiFlu™ database (https://platform.gisaid.org/epi3/cfrontend) under the accession numbers EPI_ISL_393682, EPI_ISL_393684-393703, EPI_ISL_393705-393709, EPI_ISL_393711-393723, EPI_ISL_393725-393753, EPI_ISL_393936-393946, EPI_ISL_393949, EPI_ISL_393951-393953, EPI_ISL_393955-393956, EPI_ISL_393960, EPI_ISL_393963, EPI_ISL_393965-393969, EPI_ISL_394051-394052, EPI_ISL_394107-394112.

### Collation of contemporaneous global sequence dataset

Global comparison datasets for A(H3N2) virus were retrieved from the GISAID EpiFlu™ database (https://platform.gisaid.org/epi3/cfrontend). The datasets were prepared to determine the relatedness of the viruses in this study to those circulating around the world thus understand their global context. Data processing included removal of duplicate sequences, sequences with missing dates (collection date and month), incomplete sequences, and sequences with ambiguous nucleotides (N). The data were organized into a Microsoft Excel database which also stored the associated metadata (country of origin, date of isolation, subtype, and sequence length per segment). In-house python scripts were used in the extraction and manipulation of the data. Additionally, sequences were binned by calendar year for temporal analysis. A final dataset of 1,571 global sequences sampled between January 2014 and December 2016 was available (numbers in parenthesis indicate number of sequences): Africa (281); Asia (250); Europe (250); North America (250); South America (290); and Oceania (250). The accession numbers for the global genomes are available in the study’s GitHub repository, (https://github.com/DCollinsOwuor/H3N2_Kilifi_Kenya_molecular_epidemiology_2015-16).

### Phylogenetic analysis

Consensus nucleotide sequences were aligned using MUSCLE program (https://www.ebi.ac.uk/Tools/msa/muscle/) and translated in AliView v1.26 [34]. The individual genome segments were concatenated into codon-complete genomes using SequenceMatrix v1.8 [35]. The full-length hemagglutinin (HA) sequences of all viruses were used to characterize A(H3N2) virus strains into genetic groups using Phylogenetic Clustering using Linear Integer Programming (PhyCLIP) [36] and the European CDC Guidelines [13]. Maximum-likelihood (ML) phylogenetic trees of A(H3N2) virus sequences from Kilifi and global contemporaneous sequences were reconstructed using IQTREE v2.0.7. The software initiates tree reconstruction after assessment and selection of the best model of nucleotide substitution for alignment. The ML trees were linked to various metadata in R programming software v4.0.2 and visualized using R ggtree v2.4.2 [37]. TempEst v1.5.3 was used to assess the presence of a molecular clock signal in the analyzed data, and linear regression of root-to-tip genetic distances against sampling dates were reconstructed.

### Reassortment analysis

The concatenated codon-complete genomes, based on sequences of all the eight individual gene segments of A(H3N2) viruses from Kilifi, were used to reconstruct a full-length phylogenetic tree, which was annotated by genetic group using ggtree v2.4.2 [37]. Reassortment events were assessed computationally using the Graph-incompatibility-based Reassortment Finder (GiRaF) tool [38]. The concatenated genome tree was used to infer the clustering patterns of each gene segment, show lineage classification of all eight individual gene segments, and detect reassortment events.

### Estimating A(H3N2) virus imports and exports

The ML tree topology based on Kilifi and global sequences was used to estimate the number of viral transmission events between Kilifi and the rest of the world. TreeTime [39] was used to transform the ML tree topology into a dated tree. Outlier sequences were identified by TreeTime and excluded during the process. A migration model was henceforth fitted to the resulting time-scaled phylogenetic tree from TreeTime, mapping the location status of the genomes from the nine health facilities at both the tip and internal nodes. Using the date and location annotated tree topology, the number of transitions between and within Kilifi and the rest of the world were counted and plotted using ggplot2 v3.3.3.

### Bayesian Tip-association Significance (BaTS) test

The strength of geographical clustering among the Kilifi viruses was assessed using the phylogeny-trait association test implemented in the BaTS package [40]. Each virus genome sequence was assigned a geographic code reflecting its place of sampling within the KHDSS. The overall statistical significance of geographical clustering of all Kilifi sequences was determined by calculating observed and expected association index and parsimony score statistics, where the null hypothesis is that clustering by geographical location is not more than that expected by chance. Additionally, the maximum clade statistic was used to compare the strength of clustering at each of the KHDSS location by calculating the expected and observed mean clade size from each of the nine outpatient locations. A significance level of p≤0.05 was used in all scenarios.

### Ethics

Ethical clearance for the study was granted by the KEMRI - Scientific and Ethical Review Unit (SERU# 3103) and the University of Warwick Biomedical and Scientific Research Ethics Committee (BSREC# REGO-2015-6102). Informed consent was sought and received from the study participants for the study.

## RESULTS

### IAV genome sequencing and assembly

Among the 97 IAV positive specimens that were available from the KHDSS, 72 (74.2%) that passed pre-sequencing quality control checks were loaded onto the Illumina MiSeq. A total of 63 (87.5%) codon-complete genomes were successfully assembled following sequencing: 58 (92.1%) A(H3N2) virus and 5 (7.9%) A(H1N1)pdm09 virus sequences, respectively; only the 58 A(H3N2) virus genomes were included in the analysis. The 58 genomes comprised 4 genetic groups: clade 3C.2A (n=34, 58.6%), subclade 3C.2A2 (n=3, 5.2%), subclade 3C.2A3 (n=1, 1.7%), and subgroup 3C.2A1b (n=20, 34.5%). The socio-demographic characteristics of the patients whose samples were analyzed in this report are shown in **Table 1**.

**Table 1.** Socio-demographic characteristics of the Kilifi Health and Demographic Surveillance System (KHDSS) outpatients.

|  |  | Outpatient (KHDSS) |  |
| --- | --- | --- | --- |
|  |  | n | % |
| Age | 0-11 mths | 9 | (15.52) |
|  | 12-59 mths | 18 | (31.04) |
|  | 6-15 yrs | 15 | (25.86) |
|  | 16-64 yrs | 13 | (22.41) |
|  | >=65 yrs | 3 | (5.17) |
| Sex | Female | 26 | (44.83) |
|  | Male | 32 | (55.17) |
| Cough | No | 2 | (3.45) |
|  | Yes | 56 | (96.55) |
| Difficulty breathing | No | 53 | (91.38) |
|  | Yes | 5 | (8.62) |
| Chest-wall in-drawing | No | 53 | (91.38) |
|  | Yes | 5 | (8.62) |
| Unable to feed | No | 56 | (96.55) |
|  | Yes | 2 | (3.45) |
| Oxygen saturation | <90 | 2 | (3.45) |
|  | >=90 | 56 | (96.55) |
| Conscious level | Alert / Normal | 56 | (96.55) |
|  | Lethargic | 2 | (3.45) |
|  | Prostrate | — | — |
|  | Unconscious | — | — |

### Spatiotemporal representativeness of sequenced samples

IAV was detected throughout the surveillance period in coastal Kenya (except in July, August, and September 2016) with the number of observed cases fluctuating from month-to-month (**Figure 1, Panel B**). The proportion of samples from each health facility that were sequenced roughly reflected the overall distribution of positives that were detected in the specific health facilities (**Figure 1, Panel B**). Clade 3C.2A and subgroup 3C.2A1b viruses were detected in most of the facilities (8 and 6 of the 9 health facilities, respectively), which suggests that A(H3N2) viruses were in circulation in most locations in Kilifi without geographical restriction to a particular lineage during 2015-16, (**Figure 1, Panel C**). Subclade 3C.2A2 and subclade 3C.2A3 viruses were characterized by three and one virus detection, respectively consistent with limited spread.

### Reassortment analysis

The concatenated codon-complete phylogenetic tree of Kilifi viruses revealed a topology in which the detected A(H3N2) virus genetic groups had consistent lineage classification for each of the eight gene segments, which occurs in the absence of reassortment, **Figure 2**. These findings were verified computationally using the GiRaF reassortment analysis tool, which showed no evidence of reassortment in any of the A(H3N2) viruses. The full-length codon-complete sequences, which provide a greater phylogenetic resolution, were therefore analyzed to understand the patterns of spread of A(H3N2) virus in Kilifi.

**Figure 2.**
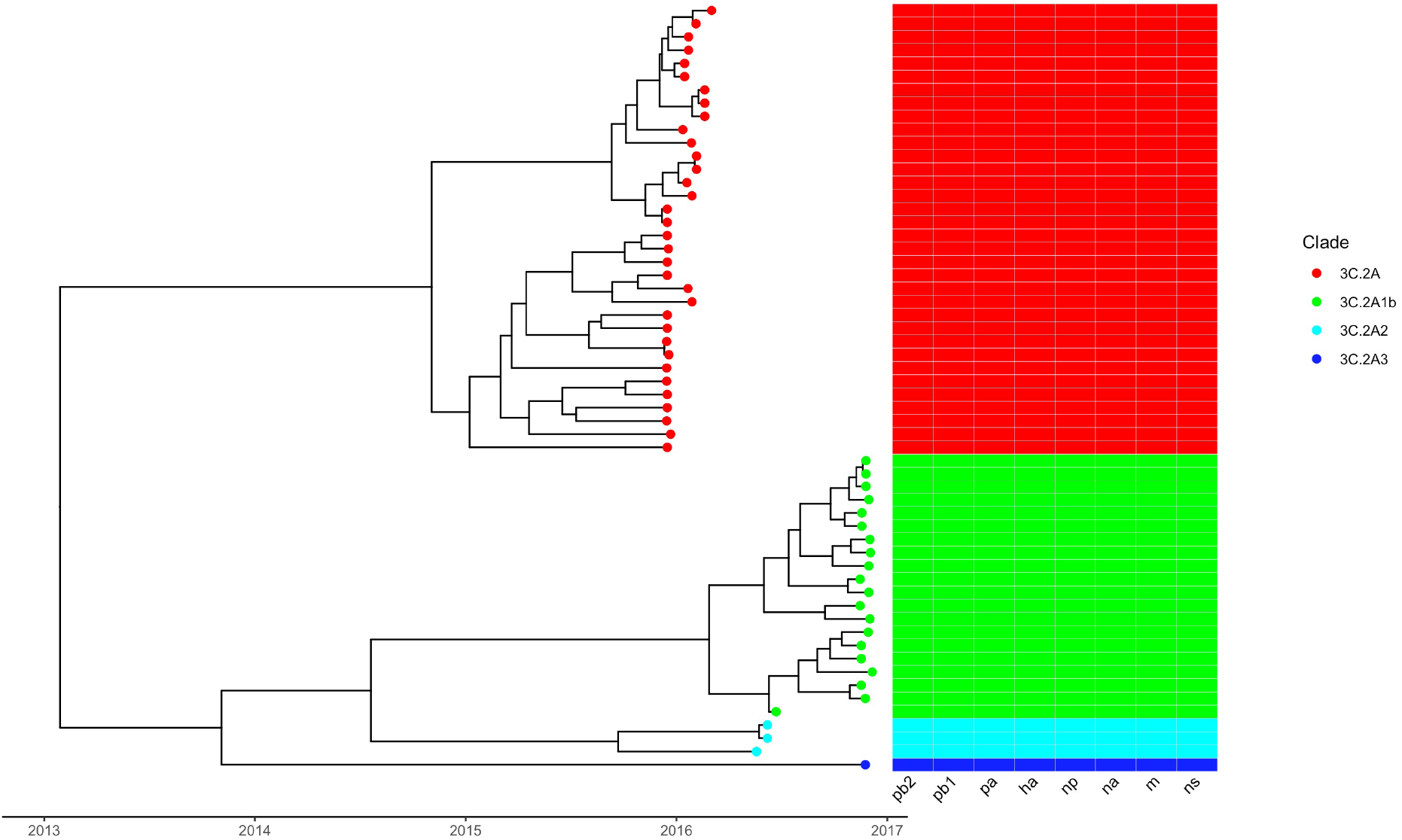
Phylogenetic tree of concatenated segments of A(H3N2) viruses from Kilifi, Kenya and schematic showing individual gene segment lineages. The time-resolved tree was constructed using 58 genome-complete sequences of A(H3N2) viruses by concatenating all the eight genome segments. Tips are colored by genetic group as follows: 3C.2A in red, 3C.2A1b in green, 3C.2A2 in cyan, and 3C.2A3 in blue. To the right is a schematic representation of viral clustering of each gene segment, which shows consistent lineage classification for all eight segments for all viruses.

### A(H3N2) virus diversity in Kilifi and viral imports and exports

We first assessed how the 58 A(H3N2) virus genomes from Kilifi compared to 1,571 genomes sampled from around the world by inferring their phylogenies. The Kilifi genomes span the existing global diversity (**Figure 3**), which suggests exchange (most likely introductions into Kilifi) of viruses with other areas around the globe. We then used ancestral location state reconstruction of the dated phylogeny (**Figure 3**) to infer the number of viral imports and exports. We inferred five importations originating from outside the region (two from Europe, two from Africa, and one from North America), which represent five independent introductions into Kilifi from areas outside the region (**Figure 4**). We also inferred a total of 23 virus location transition events between the nine health facilities, **Figure 4**. International virus imports into the study area were captured at the facilities of Chasimba (n=13, 56.5%), Matsangoni (n=6, 26.1%), Mtondia (n=2, 8.7%), and Mavueni (n=2, 8.7%), while all four virus exports from the region were captured from Chasimba facility, all occurring to Africa destinations (**Figure 4**).

**Figure 3.**
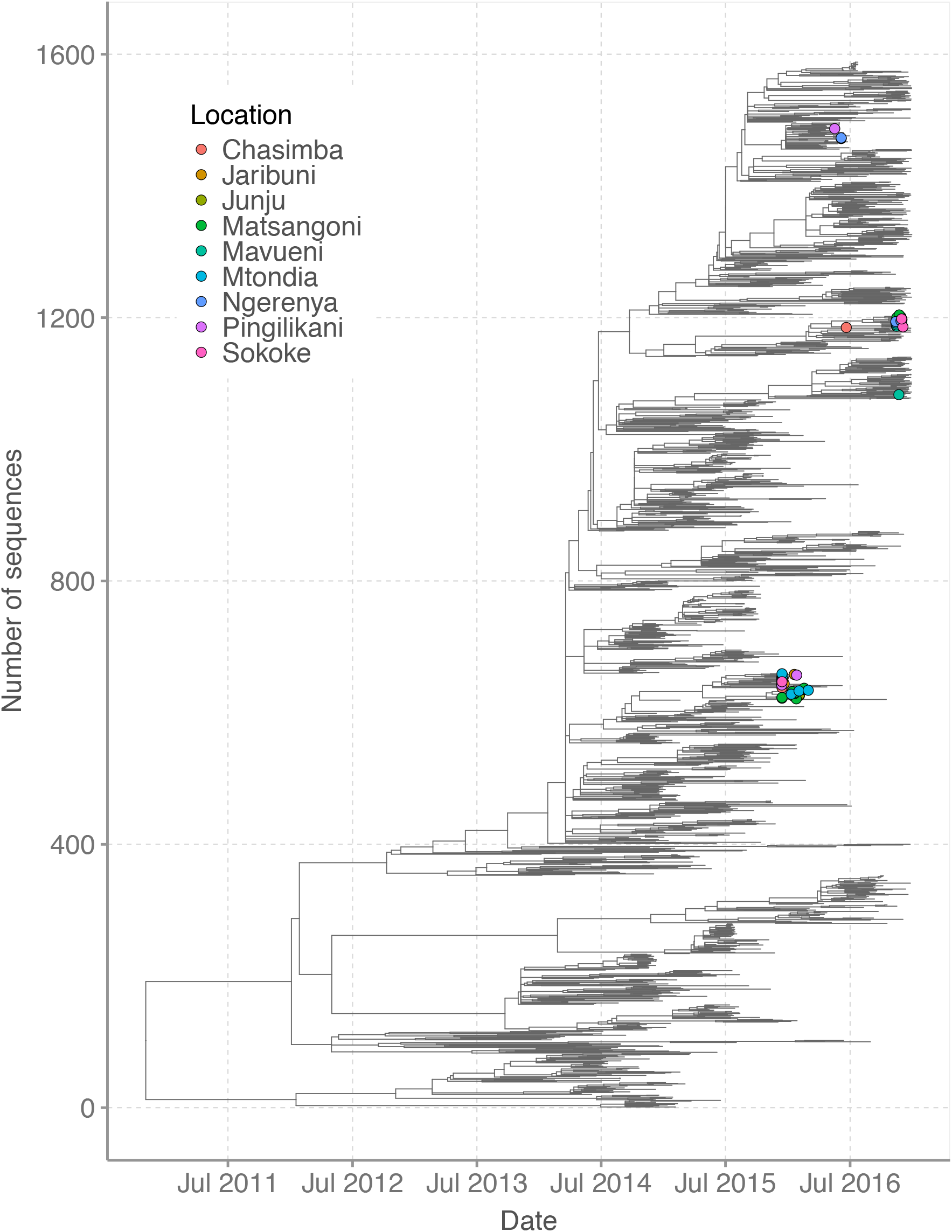
Time-resolved global phylogeny that combined 58 A(H3N2) virus sequences from Kilifi and 1,571 global reference sequences. The Kilifi sequences are indicated with filled circles colored by health facility.

**Figure 4.**
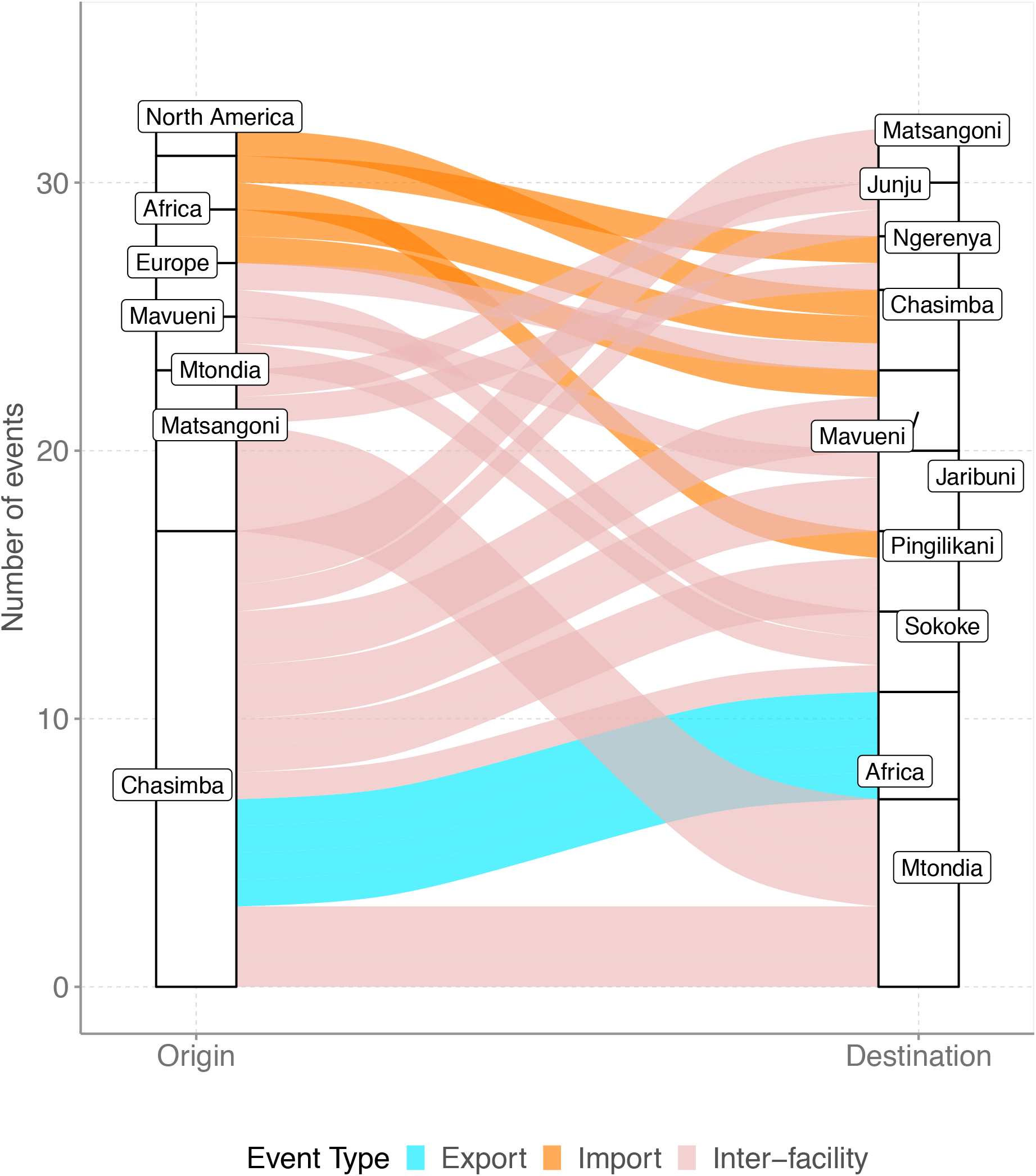
The number of viral imports and exports from Kilifi shown as alluvium plot.

### Phylogeographic structure of A(H3N2) virus in coastal Kenya

To determine the phylogeographic structure in the Kilifi sequence data using a statistical approach, phylogeny-trait association tests were conducted to determine phylogenetic association with sampling location (health facility), **Table 2**. For strains from Kilifi, the results confirmed a stronger spatial clustering of sequences at all locations (p<0.001), which is also evident in the time-resolved tree of Kilifi sequences, **Figure 5**. Additionally, the maximum clade statistic was significant (p≤0.05) in most locations (6 out of 9 locations) reflecting predominantly local evolution in most locations. The estimated differences in observed and expected maximum clade values tentatively suggested that Pingilikani and Sokoke exhibited the least spatial structure (i.e., most mixing; difference <0) while Matsangoni exhibited the strongest spatial structure (difference of 4).

**Table 2.** Bayesian Tip-association Significance (BaTS) testing results.

| Location | Association index (95% CI) ‡ |  |  | Parsimony scores (95% CI) ‡ |  |  |
| --- | --- | --- | --- | --- | --- | --- |
|  | Observed | Expected | p values | Observed | Expected | p values |
| All | 2.04 (1.77-2.32) | 5.78 (5.07-6.45) | <0.001 | 25.65 (25-26) | 37.59 (34.99-39.95) | <0.001 |

| Location | Mean maximum clade size (95% CI) ‡ |  |  |  |
| --- | --- | --- | --- | --- |
|  | Observed | Expected | p values | Difference # |
| Chasimba | 2.00 (2.00-2.00) | 1.21 (1.00-1.99) | 0.029 | 0.79 |
| Jaribuni | 1.99 (2.00-2.00) | 1.06 (1.00-1.97) | 0.004 | 0.93 |
| Junju | 3.99 (4.00-4.00) | 1.13 (1.00-1.99) | 0.009 | 2.86 |
| Matsangoni | 6.05 (6.00-7.00) | 2.07 (1.01-3.05) | 0.002 | 3.98 |
| Mavueni | 2.17 (2.00-3.00) | 1.10 (1.00-1.99) | 0.012 | 1.07 |
| Mtondia | 1.99 (2.00-2.00) | 1.46 (1.00-2.23) | 0.111 | 0.53 |
| Ngerenya | 1.99 (2.00-2.00) | 1.08 (1.00-1.99) | 0.005 | 0.91 |
| Pingilikani | 1.00 (1.00-1.00) | 1.04 (1.00-1.01) | >0.999 | -0.04 |
| Sokoke | 1.00 (1.00-1.00) | 1.04 (1.00-1.15) | >0.999 | -0.04 |

‡95% confidence interval (CI).

**Figure 5.**
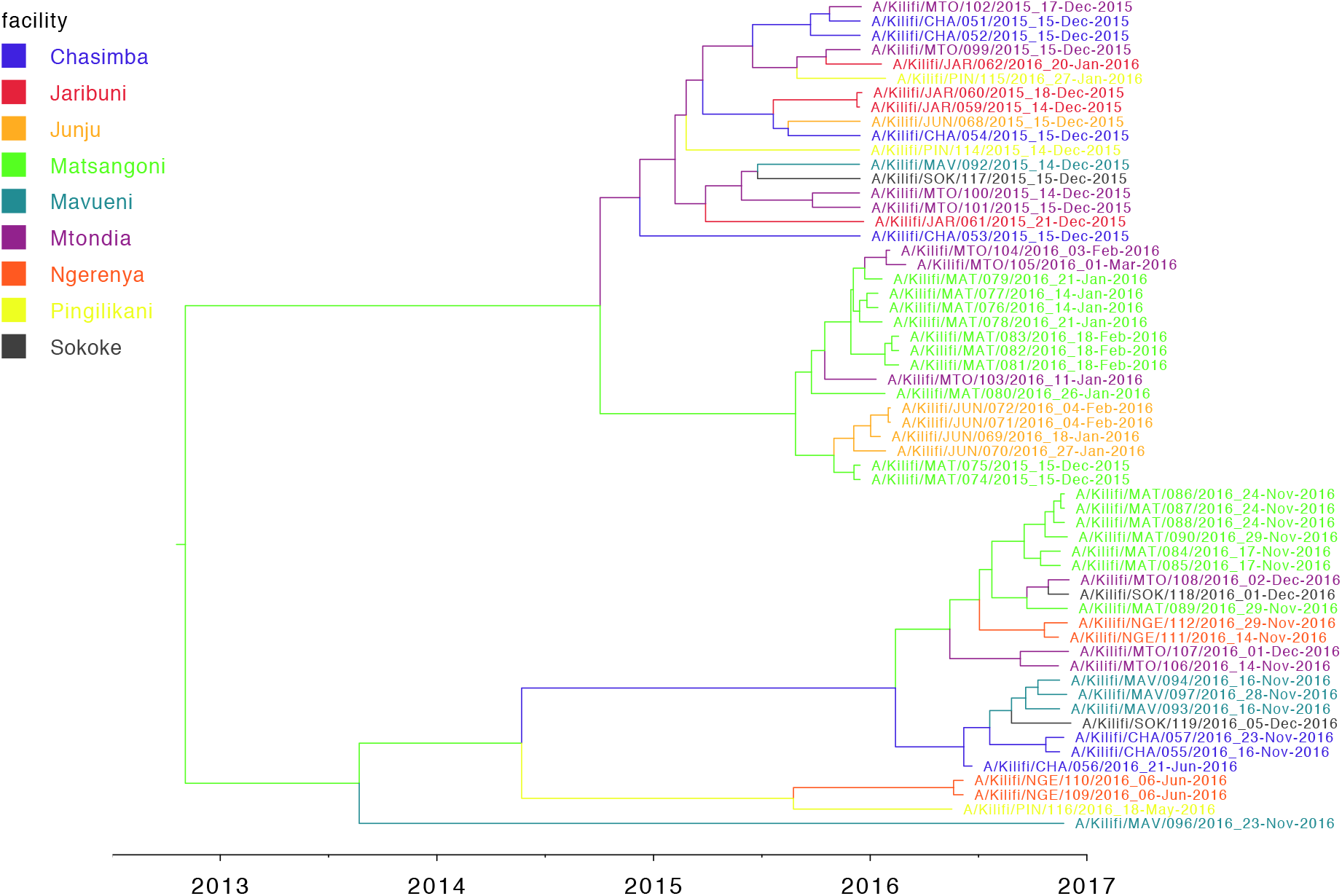
Time-resolved MCC tree inferred for A(H3N2) virus sequences from Kilifi, coastal Kenya. Branches are coloured according to the most probable location state, which is indicated in the coloured key at the top left. MCC, maximum clade credibility.

## DISCUSSION

We observed at least five independent introductions of A(H3N2) viruses into the region during the one-year period, with the importations originating from Africa, Europe, and North America. Additionally, we inferred 23 virus location transition events between the nine facilities included in the study. International virus imports into the study area were captured at the facilities of Chasimba, Matsangoni, Mtondia, and Mavueni, while all four exports from the region were captured from the Chasimba facility, all occurring to Africa destinations. We also observed a strong spatial clustering of virus strains at all locations, which was associated with local evolution.

On a global scale, annual influenza A(H3N2) virus epidemics are proposed to result from the introduction of new genetic variants from East or Southeast Asia, where viruses circulate through a network of temporally overlapping epidemics [14, 19] or from temporally migrating metapopulations of influenza viruses that seed global epidemics [18]. Here, we show that the spread of influenza A(H3N2) virus in a local community in coastal Kenya was characterized by a few virus introductions over a one-year period, followed by frequent inter-facility populations transmission. We have previously reported up to 29 IBV introductions in this population in an epidemic year predominated by IBV in 2016 [6]. This reveals that repeated introductions of A(H3N2) virus and IBV into the local population drove the influenza season in 2015-16. However, unlike the IBV epidemic, only a few introductions of A(H3N2) virus instigated the 2015-16 season, which was then sustained through local spread and evolution of circulating viruses. Therefore, the seasonal dynamics of influenza are far more complex. Wider and deeper sampling of viruses from understudied tropical and sub-tropical regions is required for a more complete understanding of the local, regional, and global spread of influenza viruses [27, 41].

The genetic diversity of A(H3N2) virus in specific regions arising from multiple virus importations and subsequent local evolution, as shown in our study, could lead to predominance of circulating viruses that are not closely matched to previously selected vaccine strains. In countries like Kenya, where virus circulation is year-round [42], there also exists unpredictability of which genetic strain may predominate and when. Influenza vaccines with broad coverage (“universal” vaccines) could be key to managing the disease burden in such settings. Currently, Kenya does not have a national influenza vaccination policy [43], but it would be important to consider deployment of quadrivalent influenza vaccines with representative A(H3N2) virus, A(H1N1)pdm09 virus, and both IBV lineages (B/Victoria and B/Yamagata) for optimal vaccine effectiveness. It will also be important to investigate further whether the use of southern hemisphere or northern hemisphere formulated vaccines could have a place in tropical regions like in Kenya, where virus importations from both hemispheres are common.

Inclusion of regional and global sequences deposited in GISAID significantly improved the power of our phylogenomic analyses, which showed that the Kilifi diversity was part of the global continuum. For example, we showed extensive geographical mixing of local strains with global strains from Europe and North America. The use of NGS technology to generate virus sequence data from Kilifi enables further scrutiny of the available data to answer other key molecular epidemiological questions. For example, the sequencing depth achieved with NGS may allow for further analysis of minority variant populations. These might be evolving locally in coastal Kenya region thus undermining the assumption that vaccines matched to globally dominant strains in trivalent/quadrivalent vaccines may necessarily protect against these local strains in a local population. Additionally, we have demonstrated the utility of NGS data analysis to investigate reassortment events in influenza viruses.

The study had some limitations. First, the study utilized samples collected from outpatients only, but would have benefitted from additional analysis of samples collected from hospitalized patients to investigate the patterns of spread of A(H3N2) virus in Kilifi. Second, the prioritized samples were selected based on anticipated probability of successful sequencing inferred from the sample’s viral load as indicated by the diagnosis Ct value. This strategy ultimately avoided NGS of some samples that may have been critical in reconstructing the patterns of spread and transmission clusters of A(H3N2) virus in coastal Kenya. Third, the KHDSS outpatient facilities surveillance collected a maximum of fifteen samples per week per site. This paucity of sequence data from some locations may have introduced bias in inference of the phylogeographic structure of A(H3N2) virus in coastal Kenya.

In conclusion, although there is paucity of studies that describe the patterns of spread of influenza on a local scale (city or town), our findings suggest that considerable influenza virus diversity circulates within defined sub-populations residing in the same geography within a country, including virus lineages that are unique to those locales, as reported for Kilifi. These lineages may be capable of dissemination to other populations through virus location transition events. Further knowledge of the viral lineages that circulate in specific locales is required to understand the main drivers of novel cases of respiratory diseases and to inform vaccination strategies within these populations.

## Data Availability

All generated sequence data were deposited in the Global Initiative on Sharing All Influenza Data (GISAID) EpiFluTM database under the accession numbers EPI_ISL_393682, EPI_ISL_393684-393703, EPI_ISL_393705-393709, EPI_ISL_393711-393723, EPI_ISL_393725-393753, EPI_ISL_393936-393946, EPI_ISL_393949, EPI_ISL_393951-393953, EPI_ISL_393955-393956, EPI_ISL_393960, EPI_ISL_393963, EPI_ISL_393965-393969, EPI_ISL_394051-394052, EPI_ISL_394107-394112.

https://github.com/DCollinsOwuor/H3N2_Kilifi_Kenya_molecular_epidemiology_2015-16

## SUPPORTING INFORMATION

### DISCLOSURE

The findings and conclusions in this manuscript are those of the authors and do not necessarily represent the official position of the Centers for Disease Control and Prevention.

### FUNDING

The authors D.C.O., F.M.N. and C.N.A. were supported by the Initiative to Develop African Research Leaders (IDeAL) through the DELTAS Africa Initiative [DEL-15-003]. The DELTAS Africa Initiative is an independent funding scheme of the African Academy of Sciences (AAS)’s Alliance for Accelerating Excellence in Science in Africa (AESA) and supported by the New Partnership for Africa’s Development Planning and Coordinating Agency (NEPAD Agency) with funding from the Wellcome Trust [107769/Z/10/Z] and the UK government. The study was also part funded by a Wellcome Trust grant [1029745] and the USA CDC grant [GH002133]. This paper is published with the permission of the Director of KEMRI.

